# Minimum Days Estimation for Reliable Dietary Intake Information: Findings from a Digital Cohort

**DOI:** 10.1101/2024.08.29.24312779

**Authors:** Rohan Singh, Mathieu Théo Eric Verest, Marcel Salathé

**Affiliations:** Digital Epidemiology Lab, School of Life Sciences, School of Computer and Communication Sciences, EPFL, Switzerland

**Keywords:** nutrition, real-world data, real-world evidence, digital cohort

## Abstract

Accurate dietary assessment is crucial for understanding diet-health relationships, but variability in daily food intake poses challenges in capturing precise data. This study leveraged data from 958 participants of the “Food & You“ digital cohort to determine the minimum number of days required for reliable dietary intake estimation. Participants tracked meals using the AI-assisted MyFoodRepo app, providing a comprehensive dataset of over 315,000 dishes across 23,335 participant days. We employed multiple analytical approaches, including Linear Mixed Models (LMM), Intraclass Correlation Coefficient (ICC), and Coefficient of Variation (CV) methods. LMM analysis revealed significant day-of-week effects, with increased energy, carbohydrate, and alcohol intake on weekends, particularly pronounced in younger individuals and those with higher BMI. ICC and CV analyses demonstrated that the required number of days varies considerably among nutrients and food groups. Water, coffee, and total food quantity by weight could be reliably estimated (ICC>0.9) with just 1-2 days of data. Most macronutrients, including carbohydrates, protein, and fat, achieved good reliability (ICC>0.75 or r=0.8) with 3-4 days of data. Micronutrients and some food groups like meat and vegetables typically required 4-5 days for highly reliable estimation. Optimal day combinations often included both weekdays and weekend days. Our findings largely align with and refine FAO recommendations, suggesting that 3-4 days, typically non-consecutive and including a weekend day, are generally sufficient for reliable estimation of energy and macronutrient intake. However, our results provide more nuanced, nutrient-specific guidelines that can inform the design of future nutritional studies.

## Introduction

Accurate dietary intake data is essential for studying the relationship between diet and health outcomes, informing public health policies, and developing effective nutritional interventions^1^. However, capturing precise dietary intake data is challenging due to the inherent individual variability in daily food consumption, recall bias, and the burden placed on participants^2^. The complexity of obtaining reliable dietary data has implications for our understanding of the role of diet on health: while many studies suggest that diet influences the risk of chronic diseases such as metabolic disorders, cardiovascular diseases, and cancer^3,4^, the strength of these associations may be limited by the quality of available dietary data^5,6^. Improving our methods for collecting accurate dietary information is therefore crucial not only for enhancing the reliability of nutritional research but also for clarifying the true extent of the impact of diet on health outcomes.

One of the primary challenges in dietary assessment is the day-to-day variation in food intake. Individuals do not generally consume the same foods in the same amounts every day, leading to variability that can obscure true dietary patterns^7–12^. For instance, Basiotis et al. showed in a small cohort of 30 people from a year long study how much variability is present, using daily food intake records for 365 consecutive days. Traditional dietary assessment methods, such as 24-hour recalls, food diaries, and food frequency questionnaires (FFQs), also have their limitations. For instance, 24-hour recalls rely on participants’ memory and may not capture infrequently consumed foods, while food diaries require significant effort from participants, potentially leading to underreporting or changes in eating behavior due to the recording process^13^.

Minimum days estimation, therefore, addresses the challenge of variability by determining the minimum number of days required to obtain a representative sample of an individual’s usual dietary intake. This approach can significantly reduce participant burden and associated costs, making dietary studies more practical and feasible^14^. By optimizing the number of days needed for accurate data collection, researchers can better allocate resources and reduce overall project costs.

Advanced dietary assessment methods, such as mobile applications for food tracking that incorporate features such as image recognition, barcode scanning, and user-friendly diet recording, offer new opportunities to refine these estimations and improve the accuracy and efficiency of dietary data collection^15,16^. These tools can facilitate personalized nutrition by providing real-world data and real-world evidence from such digital cohorts. The “Food & You” study exemplifies this, demonstrating high adherence rates to digital nutritional tracking and enabling the collection of at least two weeks of detailed dietary data from over 1,000 participants^17^.

To advance the field of dietary assessment, our study leverages the “Food & You” dataset to determine the minimum number of days required to obtain reliable dietary intake. We assessed temporal intake patterns of different food groups, macro- and micronutrients in the full cohort as well as different demographic sub-groups. Our findings indicate that while some nutrients achieve reliable estimates with as few as two days of data, others require up to a week for accurate assessments. These results have important implications for designing efficient and cost-effective dietary studies.

## Methods

### Data Collection and Preparation

The “Food & You” study involved 1,014 adults across Switzerland, with data collected from October 2018 to March 2023. The study was divided into two sub-cohorts, ‘B’ and ‘C’, where participants tracked their meals for 2 and 4 weeks respectively using the MyFoodRepo app. The MyFoodRepo app allowed study participants to track nutrition in three ways, through image taking, barcode scanning, and manual logging. Input data was then mapped to standardized nutrition tables specific to Switzerland. In particular, the images were first assessed by a machine learning algorithm using image recognition, and the result was verified, and corrected if necessary, by expert annotators specifically hired for the study. The robustness of this method is supported by both a dedicated validation study^18^ and comparisons with national dietary survey data^17^. These results indicate that the MyFoodRepo app yields data of high quality and reliability.

In the analysis of minimum days estimation below, we focused on the longest sequence of at least 7 consecutive days for each participant. This approach was chosen for several reasons: First, it allowed us to include the vast majority of participants (958 out of 1,014), with only 56 participants excluded due to insufficient data. Second, the use of consecutive days eliminated the need to account for data gaps. Third, a full week of data enabled us to investigate potential day-of-the-week effects on dietary patterns. For participants with data sequences exceeding 7 days, we calculated the mean intake per weekday to maintain consistency across the sample. The remaining 958 participants were 56% female and 44% male. Participants’ ages ranged from 18 to 65 years, with 439 participants aged below 35, and 190 participants aged above 50. BMI categories included 666 participants with a BMI between 18-25, and 281 participants with a BMI of 25 or higher. For further details on data collection and study design, see Héritier et al. (2023)^17^.

### Linear Mixed Model (LMM) Analysis

To analyze the effects of age, BMI, gender, or day of the week on nutritional intake, we employed a Linear Mixed Model (LMM) approach using the statsmodels library. This method allows for the inclusion of both fixed effects (age, BMI, gender, day of the week) and random effects (participant), accommodating the repeated measures design of the dataset. The model formula was specified as follows:

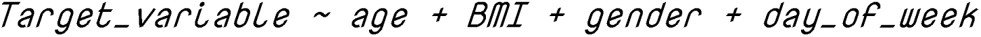

In these models, Monday was used as the reference day. From the fitted models, we extracted the model intercept, as well as the coefficients and p-values for the different days of the week. These values were then used to plot a heatmap (shown in Figure 1), illustrating the effects and significance of the day of the week on nutritional intake.

**Figure 1:**
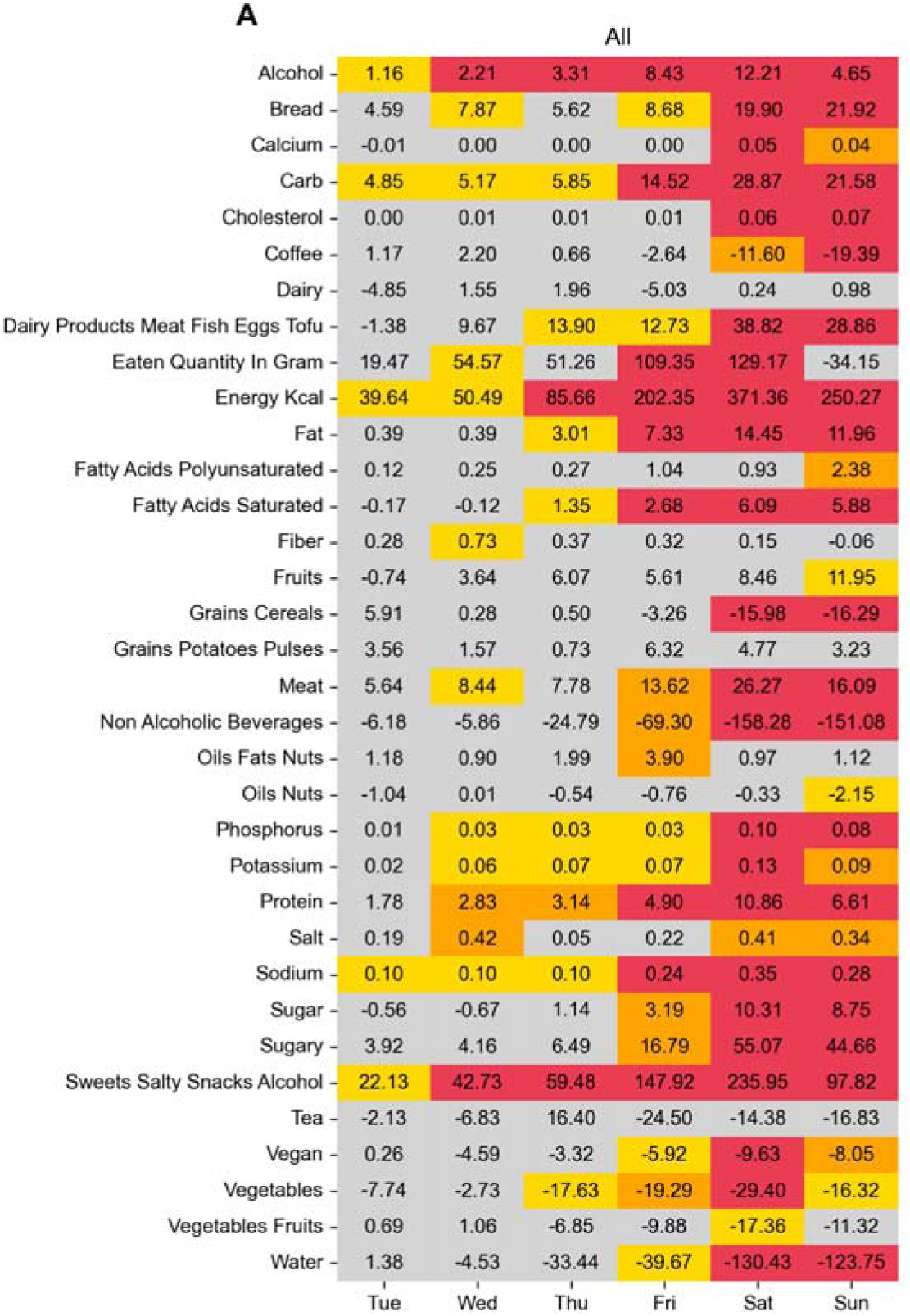
Coefficients and p values of Linear Mixed Models for various nutrients across different days of the week of all “Food & You” participants. The x-axis represents the days of the week (Tuesday to Sunday), with Monday as the reference day. The cells display the coefficients, indicating the change in nutrient intake compared to Monday. The color of the cells represents the statistical significance of the coefficients: red for p < 0.001, orange for p < 0.01, yellow for p < 0.05, and gray for p ≥ 0.05.

### ICC Calculation for Estimating Minimum Days of Dietary Assessment

Intraclass Correlation Coefficient (ICC) is a statistical measure used to assess the reliability and consistency of measurements across multiple observations. In nutritional epidemiology, ICC can be employed to determine the minimum number of days required for accurate dietary assessment. A higher ICC indicates greater reliability of the dietary intake estimate for a given number of measurement days. When examining ICC values across different numbers of measurement days, reaching a sufficiently high ICC value (typically above 0.8^19^) indicates the point at which a good estimate of an individual’s typical intake has been achieved.

We employed two distinct approaches, described below, to calculate the ICC for our dataset, using the pingouin library for ICC(3,k) calculations. The ICC(3,k) variant represents a two-way mixed effects model^19^, assessing the consistency of dietary intake across k days, thus providing an estimate of reliability for repeated measures. Our goal was to identify the point at which adding more days of dietary data collection yields diminishing returns in terms of improved accuracy.

### Approach 1: ICC Distribution Analysis

To assess the stability of ICC across different combinations of days for each nutritional feature, we computed ICC(3,k) for all possible combinations of days, ranging from k=2 to k=7. This approach yielded a distribution of ICC scores for each value of k, allowing us to examine how ICC values changed with different numbers and combinations of days. These distributions provide insights into the variability of ICC stability for a given value of k<7 across different temporal patterns within a week. A total of 38 nutritional features had at least one combination that reached the threshold of 0.75.

### Approach 2: Minimum Days and Optimal Combination Estimation

This method complements the first approach by focusing on the minimum days needed for reliable overall dietary intake estimation. We calculated the mean intake of each nutritional feature per participant across all days as a reference. We then examined subsets of days (1 to 7) in all combinations, comparing their means to the overall mean using ICC. Unlike Approach 1, which assessed consistency within subsets, this method compares subset means to the overall mean. We increased the subset size until reaching an ICC threshold of 0.9, identifying the minimum days, and the best weekday combination thereof, needed to reliably represent overall intake for a given nutritional feature.

### Minimum Days Estimation Using Variance Ratio Method

An alternative method to assess the minimum number of days required for reliable dietary assessment is the Variance Ratio Method using a linear mixed model approach. First, the model was fitted for each nutritional feature using the statsmodel library, with the day of the week as a fixed effect, and the participant as a random effect, specified by the formula:

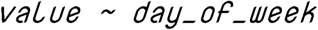

From the fitted model, we extracted the variance components, including inter-individual (σ²_b_) and intra-individual (σ²_w_) variances. These were derived from the covariance matrix of the random effects and the residual variance, respectively.

Next, we calculated the coefficients of variation for both inter-individual (CV_b_) and intra-individual (CV_w_) variances based on the extracted variances and the mean value μ of the nutritional feature. The formulas used were:

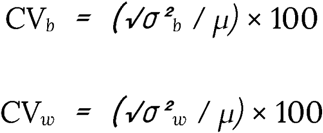

The variance ratio (VR) was then computed as the square of the intra-individual CV_w_ divided by the square of the inter-individual CV_b_, as proposed by Black et al.^10^:

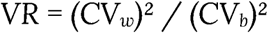

Finally, the minimum number of days (D) required to achieve specified reliability thresholds r = 0.8, 0.85, 0.9 (‘r’ is the expected correlation coefficient between observed and true intakes), was calculated using the formula: D = (r² / (1 - r²)) × VR. This process was applied to each nutritional feature for the aforementioned reliability thresholds, yielding the minimum number of days required for reliable dietary assessment.

## Results

### Consumption patterns across days of the week

It has been documented that nutrient intake can vary significantly across different days of the week^20–22^. To analyze the association of different days of the week and nutrient intake, we used the linear mixed models approach described in the methods section. The results are plotted as a heatmap (Figure 1) where the p-values indicate the statistical significance of the coefficients, with thresholds set at <0.001 (red), <0.01 (orange), <0.05 (yellow), and ≥0.05 (gray). The numerical values within the cells represent the coefficients of the day of the week for each nutrient, indicating the change in nutrient intake compared to the intercept (with Monday as reference). Furthermore, the results are segmented into groups as follows: all participants (Figure 1), age groups (Supplementary Figure 1), BMI groups (Supplementary Figure 2), and gender groups (Supplementary Figure 3) in order to observe the variations of dietary daily habits with respect to age and BMI.

We observed significant variations across different days for several nutrients and food groups, as shown in Figure 1. Alcohol intake shows notable increases from Wednesday through Sunday (all p < 0.001). Similarly, carbohydrate intake increases significantly throughout the week, peaking on Friday, Saturday, and Sunday (increase of 15g, 29g and 22g respectively; p < 0.001). Energy intake followed a similar pattern, with substantial increases on all days, particularly on Friday, Saturday, and Sunday (+202 kcal, +371 kcal, and +250 kcal respectively, p < 0.001). The intake of sodium and sugar also increased significantly on the weekend, with sugar increasing by roughly 10g on Saturday and Sunday, respectively (p < 0.001). These observations reflect more indulgent dietary habits towards the end of the week. In contrast, non-alcoholic beverages, especially water and coffee, exhibited a significant decrease from Friday to Sunday (p < 0.001). Notably, dairy was the only food group for which there was no significant difference across all days of the week, even when considering subgroups (Supplementary Figures 1-3).

For individuals aged 35 years or younger (Supplementary Figure 1A), carbohydrate consumption pattern was similar to the overall study population, with significant increases on Thursday, Friday, and Sunday (+15g, +30g and +18g respectively, p < 0.001). Interestingly, these significant increments were also seen for other weekdays for the older age group, with an increase of about 11g from Tuesday to Thursday (p < 0.05), shown in Supplementary Figure 1B. Eaten quantity for the younger age group changed significantly over the weekend with the exception of Sunday where it decreased (-90g, p < 0.01). In the age group 50+, this increase was not significant for any other days. Fruit consumption significantly increased during the weekend (between 35-40g) as well among the older age participants. Note that fruit consumption in this study comprises consumption of both fruit juices and whole fruits, which might explain why the younger age group displayed a much higher baseline for this food category. Converse to the previous nutrient trends, intake of non-alcoholic beverages such as coffee and water decreased significantly, particularly during the weekend (all p < 0.001) in the younger age group, while this difference was either non-significant (coffee, tea and non-alcoholic beverages) or not as significant (water, p < 0.05) in the older age group. Vegetable consumption on Saturday and Sunday also decreased (-33g and -28g, p < 0.01) significantly in the younger age group but non-significantly in the older age group.

For individuals with a healthy BMI (between 18 and 25), carbohydrate and energy intake increased significantly from Friday onwards to Sunday (all p < 0.001), as highlighted in Supplementary Figure 2A, akin to the observation for the full cohort. Interestingly, the intake of meat, and thus also protein consumption, steadily increased significantly over the week. Similar to the full cohort, intake of non-alcoholic beverages, including coffee and water, showed a notable decrease on the weekend (p < 0.001), while vegetable intake decreased significantly on Saturday (- 26g, p < 0.01). For individuals with a BMI greater than 25 (Supplementary Figure 2B), carbohydrate and energy intake patterns are similar to those with healthy BMI. Conversely, meat consumption was higher among those with high BMI, but its consumption over the week was not significantly higher than the baseline with the exception of Saturday. The consumption of vegetables and grains-cereals decrease was more pronounced over the week among high BMI participants. For potassium intake, the increment was observably the highest on Saturday in both the groups (p < 0.001 for healthy BMI; p < 0.05 for high BMI), while for sodium, the intake was substantially higher on the weekend (Friday to Sunday with p < 0.001) for the healthy BMI group. Sodium intake among high BMI participants was also higher during the weekend, substantially higher on Saturday (p < 0.001), but slightly lower on Sunday (p < 0.01) and Friday (p < 0.05). Finally, when comparing consumption patterns by gender, males had higher increases in consumption of meat, daily eaten quantity, and kcal energy across several days of the week compared to females (Supplementary Figure 3), consistent with findings from earlier studies^23,24^.

When using mixed models to analyze dietary data, certain limitations may arise that suggest the need for complementary methods like ICC and CV. Mixed models estimate population-averaged effects, which can overlook subtle day-to-day variations within individuals. Additionally, skewed or non-normal data distributions can impact the model’s accuracy, and assumptions about random effects might obscure real patterns. Due to these potential limitations, using ICC and/or CV based approaches may help in capturing within-subject variability and providing a clearer picture of daily dietary behaviors.

### Observed ICC Patterns in Nutrient Features

The ICC values for various nutrient features were computed over different numbers of days to determine the optimal duration of dietary assessment for reliable estimates of average nutrient consumption. Figure 2 shows the top 24 distinct food features that reached an ICC threshold of 0.75 - the remaining features are shown in Supplementary Figure 4. The x-axis in both figures represent the number of days, while each boxplot demarcates the ICC scores for combinations of days for each minimum number of days on the x-axis. The ICC thresholds of 0.9 and 0.75 are shown as a horizontal red line in each subplot. The color of the boxplots represents different nutrient groups: blue for micronutrients, red for food groups, and green for macronutrients. The ICC values for all nutrients generally improve as the number of days of dietary data increases. This indicates that more extended periods of dietary assessment tend to provide more reliable estimates, as expected. Largest daywise variability for all nutrients occurs when only 2 days of data are used.

**Figure 2:**
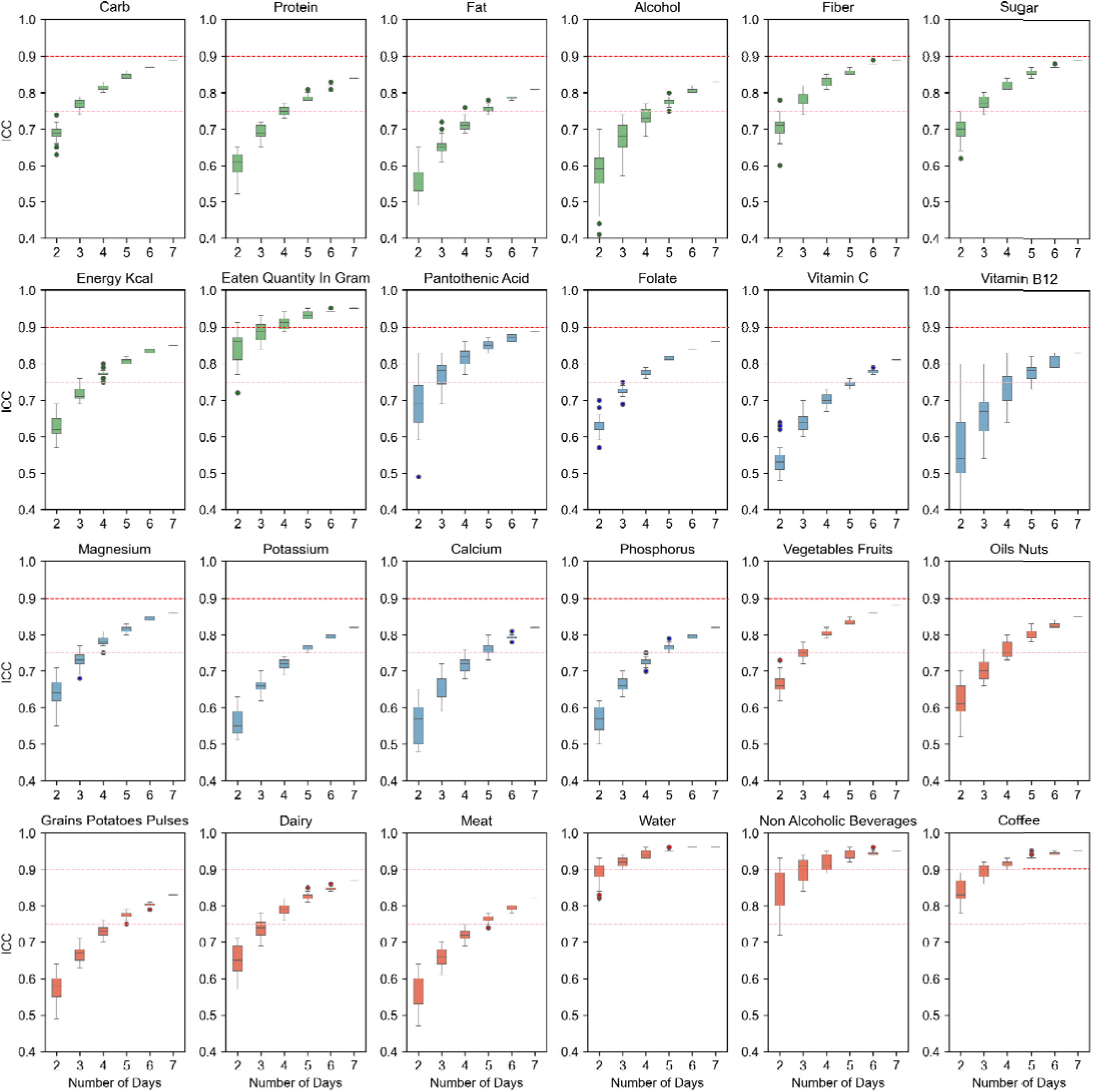
Intraclass Correlation Coefficient (ICC) values for various nutrient intake measurements across different numbers of days. The x-axis represents the number of days, while the y-axi represents the ICC scores. Each subplot corresponds to a different nutrient, with boxplot indicating the ICC scores for combinations of days for each minimum number of days on the x-axis.

It took 3 days for the median ICC of carbohydrate intake to attain a value of 0.75, which is widely considered the minimum threshold for good reliability^19^. For protein and fat intake, it took longer, about 4-5 days, to attain this threshold. Similarly, the median ICC for alcohol intake required 5 days, while sugar and fiber intakes required just 2-3 days to reach 0.75, and in about 7 days nearly reached the ICC threshold of 0.9, which is considered excellent reliability. Four days was sufficient for the median ICC of energy kcal consumption to reach the 0.75 threshold, while this was just 2 days for eaten quantity. Micronutrients, like magnesium, pantothenic acid, folate, phosphorus and calcium, were able to cross the 0.75 threshold at 4-5 days, as observed by their median ICCs from their day-wise combinations for these days. Interestingly, water and non-alcoholic beverages intakes cross the 0.75 threshold in just 2 days. Many combinations of food groups, like vegetables-fruits and oil-nuts, as well as dairy, attained the 0.75 threshold at 4 days, while for meat and grains-potatoes-pulses, reaching the 0.75 threshold required at least 5 days.

In summary, using this method, most nutrients show a stabilization of ICC values above the 0.75 threshold after 4-5 days, suggesting that to achieve high reliability for most nutrient intake measurements a 4-5 days of dietary logging is desirable. Certain nutrients, typically non-alcoholic drinks such as water, achieve higher ICC values even with fewer days of measurement, indicating that these nutrients may require less extensive data collection to reach reliable estimates.

The horizontal red lines in each subplot denote the ICC threshold of 0.75 (good reliability) and 0.9 (excellent reliability). Boxplot colors represent different nutrient groups: blue for micronutrients, red for food groups, and green for macronutrients. Note that ICC does not generally reach 1 due to inherent within-subject variability.

### Minimum Days and Optimal Combination Estimation

In order to extract the best (and worst) collection day combinations for each nutrient or food group, we used another approach wherein we compare the ICC of different day combinations to the mean intake of the entire dataset (consisting of a 7 day week). Hence, under the assumption of an entire week being sufficient in capturing the nutritional variation, this method allows to elucidate which days of the week (or their combinations) lead to highest (or lowest) ICC values when comparing to the full week.

Figure 3 highlights the best and worst day combinations at different number of days across various nutritional features (for the full set of nutrition features, see Supplementary Figures 5-6). Since the assumption of one week being sufficient to capture variation already loses some precision, we set the ICC threshold in the assessment below at 0.9, indicating excellent reliability. In order to achieve this threshold, carbohydrate intake required 3 days that were spread across the week (Monday, Wednesday and Saturday) for the best combination. The worst combination of 3 days also reached the 0.9 threshold, but the days were continuous weekdays from Monday to Wednesday. Similarly, for protein and fat intake, it required 3 days minimum for the best combination to cross the 0.9 threshold and included two weekdays and one weekend day (Saturday for both protein and fat), while their worst 3 day combination was below the 0.9 threshold. For fiber, the best combination of 2 days (Monday and Wednesday) attained the 0.9 threshold precisely, although with 3 days, even the worst combination was above the excellent reliability threshold. For alcohol, the alternating day combination at 3 days resulted in being the best combination which surpassed the threshold, while the worst combination at even 4 days - which were all continuous weekdays - was below the threshold. Eaten quantity and water intake required only one day of collection (Wednesday) to reach the reliability threshold. The same can be observed for non-alcoholic beverages group (Supplementary Figure 5) Coffee intake nearly achieved this threshold with just one day of data (Tuesday). Interestingly, when adopting a more conservative approach by increasing the collection period to 2 days, the optimal combination for all four nutritional features (eaten quantity, water, non-alcoholic beverages and coffee) was consistently Monday and Friday. Similarly, extending to a 3 day collection period, the best combination was Monday, Tuesday, and Friday for all these features (except for non-alcoholic beverages).

**Figure 3:**
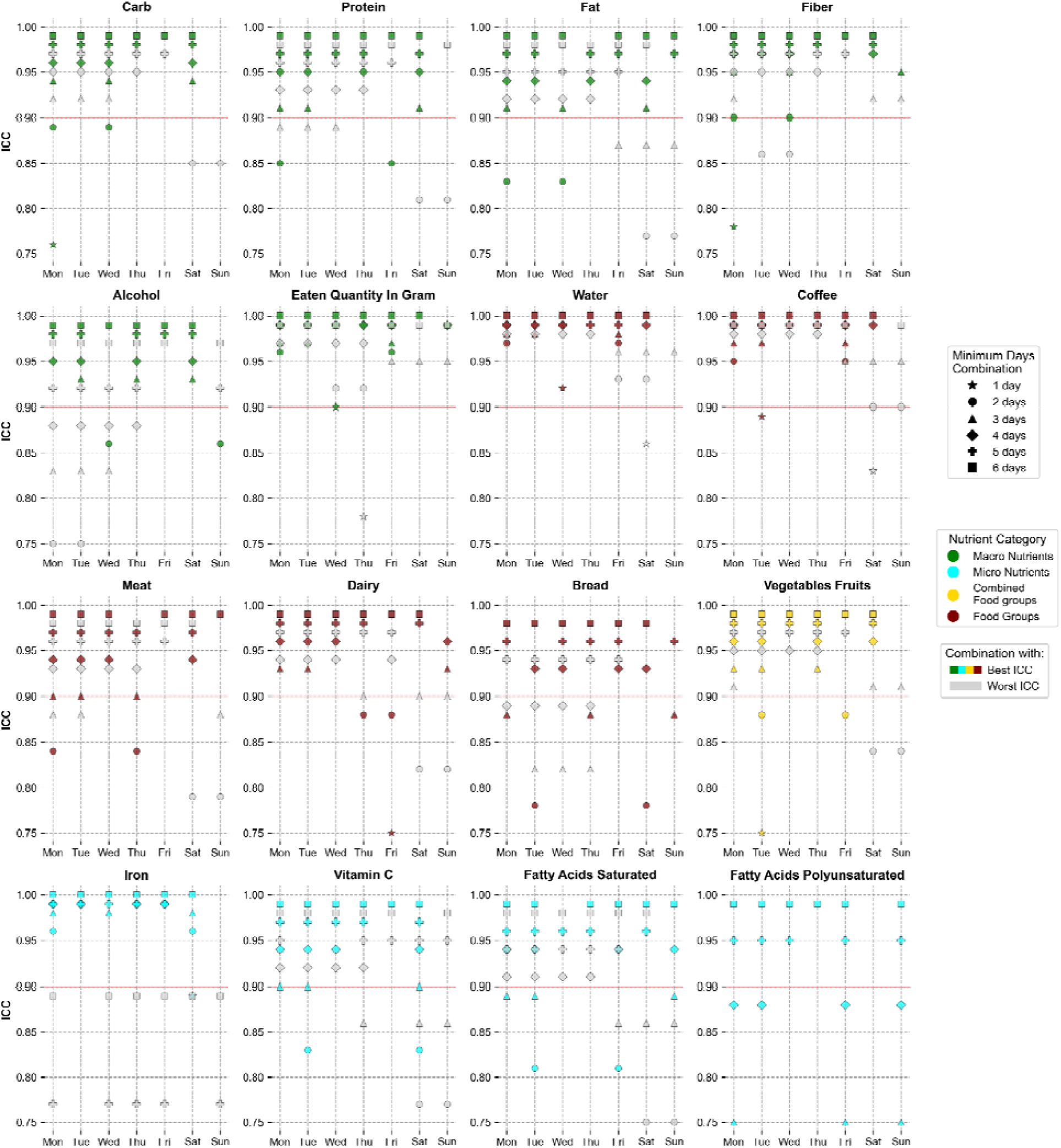
Best and worst day combinations at different minimum days for reliable dietary assessment of different nutrients / food groups. For each nutrient and at each number of days, the day combinations which yielded the highest (in color) and lowest ICC scores (in gray) are shown. The plot is ranged between ICC values of 0.75 to 1.0, with the ICC reliability threshold at 0.9 shown as a red line - points lying below this range are not shown.

A 3-day combination of Monday, Tuesday, and Thursday precisely reached the 0.9 reliability threshold for meat intake. Notably, when extending the collection period to 4 days, even the least optimal combination - consisting of four consecutive days - surpassed this threshold. This suggests that any 4-day period within a week is sufficient to accurately capture an individual’s meat consumption patterns. For dairy intake, the least optimal combination at 3 days, consisting of two weekend days, had exactly reached the 0.9 threshold, while the best combination consisted of Monday, Tuesday and Sunday. Bread consumption needed 4 days for its best combination to surpass the 0.9 threshold. The collective vegetables and fruit group required just 3 days for the worst day combination to reach the threshold, while the best combination included only weekdays (Monday, Tuesday and Thursday).

Interestingly, the optimal 2-day combination for iron intake (Monday and Saturday) exceeded the 0.9 threshold. However, unlike other features, even some 6-day combinations that included both Monday and Saturday fell below this threshold. This unexpected result suggests that the observed patterns in iron intake may be subject to high variability or influenced by factors not fully captured in our data. For vitamin C, the best combination at 3 days (Monday, Tuesday and Saturday) reached the 0.9 threshold exactly, although a more conservative choice of 4 days yields ICC values above 0.9 even for its least optimal combination. Other vitamins like B1, B2, B6 and D required at least 4-5 days for the most optimal combination to cross the threshold, while vitamin B12 required just 3 days (Supplementary Figure 6). For saturated fatty acids, the results were similar to vitamin C, with 4 days yielding ICC values above 0.9 even for the least optimal combination. Polyunsaturated fatty acids and monounsaturated fatty acids, however, required more days (Figure 3 and Supplementary Figure 6 respectively): only the best 5-day combination crossed the 0.9 threshold, while some 5-day combinations produced ICC values below 0.75, highlighting greater variability in intake.

### Minimum days estimation covariance method

In addition to the Intraclass Correlation Coefficient (ICC) method, we also employed the Coefficient of Variation (CV) method to estimate the minimum number of days required for reliable dietary assessment. While both methods account for within-person and between-person variability, the CV method, as demonstrated by Black et al.^10^, provides a direct calculation of the number of days needed to achieve specific reliability thresholds. The CV method uses the ratio of within-person to between-person variation to calculate the number of days required to achieve a specified correlation (r) between observed and true mean nutrient intake (see Methods), with higher values indicating greater reliability.

Figure 4 shows the minimum number of days required to achieve high to very high reliability thresholds of 0.8 (blue), 0.85 (yellow), and 0.9 (green) for various nutrients and food groups. Similar to the observations from the ICC approach (Figure 3 and Supplementary Figure 4), water, coffee, non-alcoholic beverages, and eaten quantity by weight can be reliably estimated (at r=0.85) with just 1 day of data collection. Furthermore, with the exception of the eaten quantity feature, we observed for the other three features that their between-subject coefficient of variations (CV_b_) were quite a bit larger compared to their within-subject coefficient of variations (CV_w_), thereby reducing their variance ratios. This indicates that individuals have very consistent patterns in their fluid intakes and overall food quantity consumption, but simultaneously, these intakes vary across individuals.

**Figure 4:**
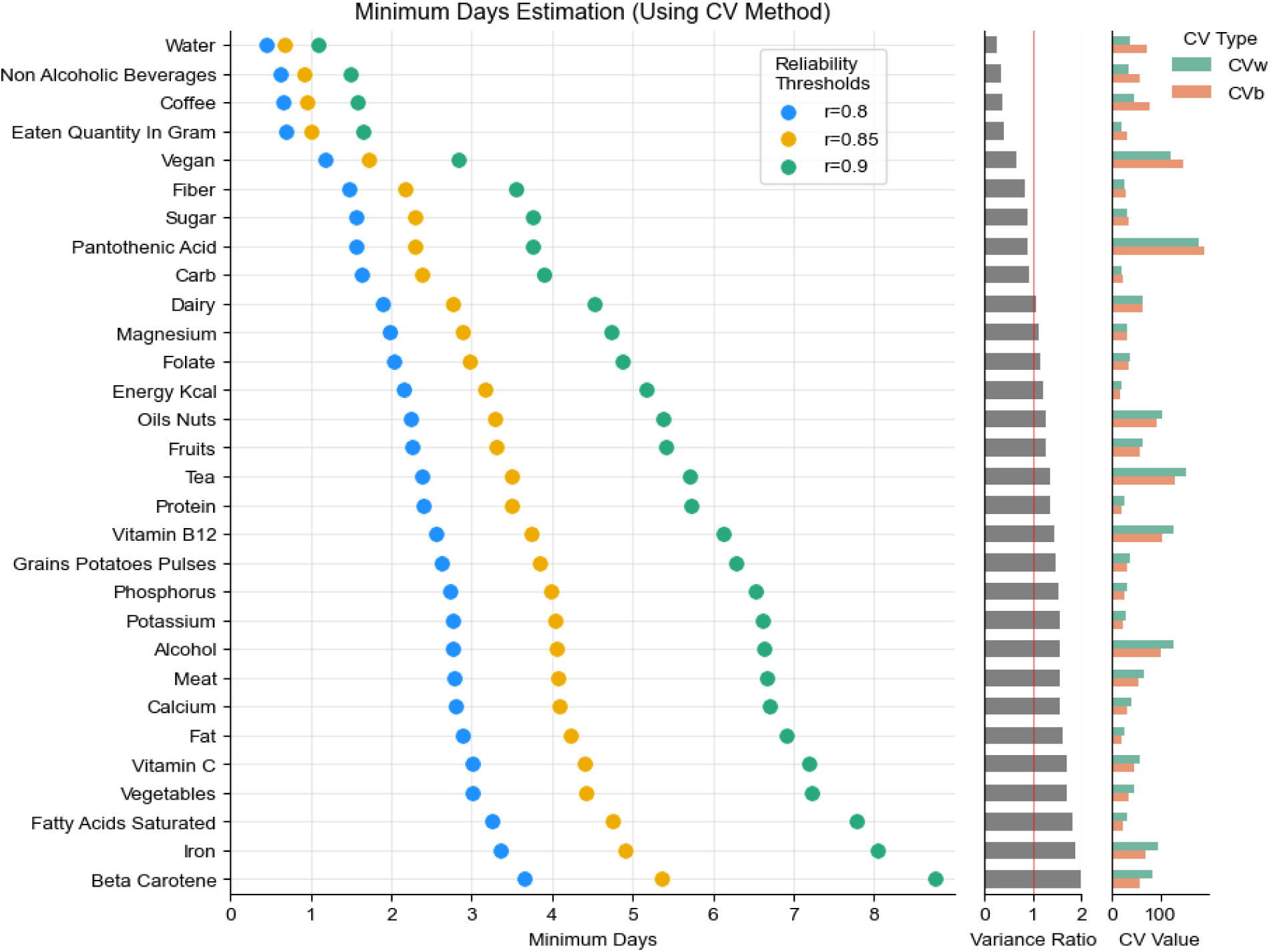
Minimum number of days required to achieve reliability thresholds (r) of 0.8 (blue), 0.85 (yellow), and 0.9 (green) for various nutrients and food groups using the Coefficient of Variation (CV) method. These thresholds represent different levels of desired correlation between observed and true intakes, with 0.9 being the most stringent criterion. Top 30 nutrients are shown; sorted by the variance ratio. Additionally, variance ratio, between-subject coefficient of variation (CV_b_), and within-subject coefficient of variation (CV_w_) are shown for the top 35 nutrients.

Similarly, major macronutrient intake like carbohydrates, sugar, fiber, fat, and protein as well as energy consumption (measured in kilocalories), also showed high reliability with only 2-3 days of data (at r=0.8). Micronutrients, on the other hand, tended to require longer periods for reliable estimation. Saturated fatty acid, beta carotene, vitamins C as well as certain minerals like phosphorus, potassium, calcium and iron typically needed 3-4 days of data (at r=0.8 threshold) to achieve decent reliability. Moreover, iron intake also displayed high CV_w_, which may substantiate the aberrant ICC minimum days estimation in Figure 3. However, some micronutrients like folate and magnesium required only 2 days. Food groups such as dairy and fruits needed only 2 days for high reliability (r=0.8), and about 5 days for very high reliability (r=0.9). Meat and vegetables required longer, about 3 days for high reliability, and correspondingly longer - 7 days - for very high reliability.

## Discussion

Recent advances in digital dietary assessment tools offer promising opportunities to enhance data collection in nutritional epidemiology. For instance, the “Food & You” study, consisting of over 1,000 participants who tracked their dietary intake using the AI-assisted MyFoodRepo app, used such a digital approach to capture detailed and accurate dietary intake in a real-world setting^17^. However, despite the advantages of such technologies, it is still crucial to determine the minimum number of days required for reliable dietary intake estimation, in order to design nutritional studies and extract meaningful individual dietary profiles. Optimizing the duration of dietary assessment offers multiple benefits: it can reduce participant fatigue and dropout rates while simultaneously improving the cost-effectiveness and feasibility of large-scale nutritional studies. To determine the ideal duration of data collection, a sufficiently powered study with long-term data is needed. This allows for systematic analysis of how reducing data collection periods affects the reliability of dietary intake estimates. While other studies may have also collected extensive dietary data, our analysis, based on the Food & You dataset, represents one of the most comprehensive examinations of this specific question to date. The Food & You study encompasses more than 46 million kcal collected from 315,126 dishes over 23,335 participant days^17^, providing a robust foundation for our investigation. The current study thus offers unique quantitative measures that can guide study planning across various levels of nutrient information, from micronutrients to broad food groups, and at different reliability thresholds.

Our Linear Mixed Model (LMM) analysis revealed distinct weekly patterns in nutrient intake, highlighting the variability of consumption profiles both across and within individuals. Significant increases in energy, carbohydrate, and alcohol intake were observed on weekends, particularly on Saturdays, indicating more indulgent dietary habits during these periods. For instance, carbohydrate intake increased by 29g on Saturdays (p < 0.001) compared to the Monday baseline, while energy intake rose by 371 kcal (p < 0.001). Concurrently, we observed significant decreases in the consumption of non-alcoholic beverages, especially water and coffee, from Friday to Sunday (p < 0.001). These patterns were more pronounced in younger individuals (aged 35 or below) and those with higher BMI. Gender differences were also apparent, with males exhibiting higher increases in meat consumption and overall eaten quantity across several weekdays compared to females. These findings underscore the importance of accounting for day-of-week effects, as well as demographic factors, in dietary assessments and interventions.

To determine the minimum number of days required for reliable dietary intake estimation, we first used the Intraclass Correlation Coefficient (ICC) method. Our results indicate that the required number of days varies considerably among different nutrients and food groups. For macronutrients, carbohydrate intake achieved good reliability (ICC > 0.75) with 3 days of data, while protein and fat intake required 4-5 days to reach the same threshold (Figure 2). Our analysis of optimal day combinations revealed that spreading these days across the week, including both weekdays and weekend days, generally yielded higher reliability (Figure 3). For instance, the best 3-day combination for carbohydrates included Monday, Wednesday, and Saturday, which precisely reached the excellent reliability threshold (ICC = 0.9). Interestingly, some dietary components, such as water, non-alcoholic beverages, coffee, and total daily eaten amount, could be reliably estimated (ICC > 0.9) with as little as 1-2 days of data. In contrast, food groups like meat, vegetables, and alcohol required at least 3-4 days for good reliability. Meat intake, for example, reached the 0.9 reliability threshold with a 3-day combination of Monday, Tuesday, and Thursday. Micronutrients typically exhibited more variability, necessitating longer periods of data collection. For instance, magnesium, pantothenic acid, folate, phosphorus, and calcium required 4-5 days to cross the 0.75 ICC threshold.

To complement and validate our ICC findings, we also employed the Coefficient of Variation (CV) method to estimate the minimum number of days required for reliable dietary assessment. The CV method, which directly calculates the number of days needed to achieve specific reliability thresholds, largely corroborated our ICC results while providing additional insights. Consistent with our ICC analysis, the CV method indicated that water, coffee, non-alcoholic beverages, and eaten quantity by weight could be reliably estimated (r=0.85) with just one day of data collection (Figure 4). This alignment between methods reinforces the robustness of these particular findings. Similarly, both approaches suggested that major macronutrients (carbohydrates, sugar, fiber, fat, and protein) and energy consumption required only 2-3 days of data for high reliability (r=0.8 in the CV method, comparable to ICC>0.75). The CV method also confirmed our ICC findings regarding micronutrients, showing they generally required longer periods for reliable estimation. For instance, saturated fatty acids, beta carotene, vitamin C, and minerals like phosphorus, potassium, calcium, and iron typically needed 3-4 days of data to achieve good reliability (r=0.8).

We also benchmarked our minimum days observations from the covariance (CV) method against two comparable studies, Pereira et al. (2010)^8^ and Palaniappan et al. (2003)^9^, which also employed the same method for several macro- and micro-nutrients. Several of these nutrients showed notable similarities for minimum days. For instance energy intake, the minimum days in our study was 2 days (at r=0.8), which closely matches with Pereira et al. for both males and females, who reported 3 days each (at r=0.8). This also aligns reasonably with Palaniappan et al., which found 2 days for men and 4 days for women (at r=0.8). Similarly, for carbohydrate intake, our study yielded 2 days at r=0.8, which is strikingly consistent to both other studies at 2-3 days. Fiber intake also showed some alignment, with our study indicating 2 days and 3-4 days for Pereira et al. Iron intake also showed strong agreement, with our study indicating 3-5 days (r=0.8 and r=0.85 respectively), while Pereira et al. found 4 days for females, and Palaniappan et al. reported 4 days for both men and women. For calcium, both our study and Pereira et al. reported 4 days (for both males and females), while just 2 days by Palaniappan et al. Saturated fat shows a strong match as well, with our study indicating 5 days and Pereira et al. reporting the same for females. Cholesterol intake in our study was 6 days, aligning closely with Pereira et al.’s finding of 7 days for females. Protein intake, however, was quite dissimilar across the three studies, wherein we report ∼3 days, while Pereira et al. reported 8 days for males and 4 days for females, and Palaniappan et al. indicated 3 days for men and 6 for women. This was also the case for total fat consumption, where we found the minimum days to be 3-4 days (at r=0.8 and r=0.85 respectively), while ranging between 3-7 days in Pereira et al. and 2-5 days in Palaniappan et al.’s study.

Our findings largely align with and refine the recommendations set forth by the Food and Agriculture Organization (FAO)^25^, which suggests that a minimum of three to four days of dietary intake data, including a weekend day, is generally required to accurately characterize an individual’s usual intake of energy and macronutrients. Indeed, our results from both the ICC and CV methods corroborate this guideline, indicating that for most macronutrients and energy intake, 3-4 days of data collection yield reliable estimates. Importantly, our analysis of day-of-week effects and optimal day combinations strongly supports the FAO’s emphasis on including weekend days, as we found significant variations in intake patterns between weekdays and weekends for many nutrients. However, our study provides a more nuanced understanding by demonstrating that the required number of days can vary significantly depending on the specific nutrient or food group of interest. For instance, while some dietary components like water and coffee intake can be reliably estimated with just 1-2 days of data, others, particularly micronutrients and certain food groups, may require up to 5-7 days for very high reliability. These insights offer more precise guidelines for designing dietary assessment protocols.

While our study provides valuable insights into the minimum days required for reliable dietary assessment, several limitations should be acknowledged. Firstly, our analysis is based on a single cohort study conducted exclusively in Switzerland. As such, the findings may not be fully generalizable to other populations or cultural contexts with different dietary patterns and habits. Additionally, while the “Food & You” study included over 1,000 participants, its sampling methods did not yield a fully representative population sample, which may limit the broader applicability of our results. Furthermore, although the MyFoodRepo app collects accurate dietary data, it’s important to note that the information is still self-reported, which introduces potential biases inherent to this method of data collection. While all the collected data was verified by expert human annotators, challenges like selective reporting or changes in eating behavior due to the act of recording remain possible. Lastly, while our study duration allowed for the analysis of weekly patterns, longer-term studies might reveal additional insights into dietary variability over extended periods. Given these limitations, further research is needed to validate our findings across diverse populations, cultural settings, and longer time frames. Future studies should also aim to compare digital dietary assessment tools like MyFoodRepo with other methods to further establish their reliability and potential advantages in nutritional research.

In conclusion, our study provides insights into the optimal duration of dietary data collection, offering a nuanced understanding that can enhance the design and implementation of nutritional research. Determining the minimum number of days required for reliable dietary intake assessment across various nutrients and food groups may enable more efficient, cost-effective, and participant-friendly dietary studies. These findings have implications for both personalized nutrition approaches and broader public health policies. The variability we observed in data collection requirements across different nutrients underscores the importance of tailoring study designs to specific research questions or interventions. Moreover, our results highlight the value of leveraging real-world data and real-world evidence from digital cohorts, demonstrating how tools like AI-assisted food tracking apps such as MyFoodRepo can contribute to more accurate and comprehensive dietary assessments. As we move towards an era of precision nutrition, these insights could be important for developing more targeted and effective dietary interventions, ultimately contributing to improved health outcomes at both individual and population levels. Future research should build upon these findings, exploring their applicability across diverse populations and cultural contexts, and further investigating the potential of digital tools in nutritional epidemiology.

## Data Availability

All data produced in the present study are available upon reasonable request to the authors

## Data Availability

The data that support the findings of this study are not publicly available due to privacy. However, data can be made available upon request from the corresponding author.

---

**Supplementary Figure 1:**
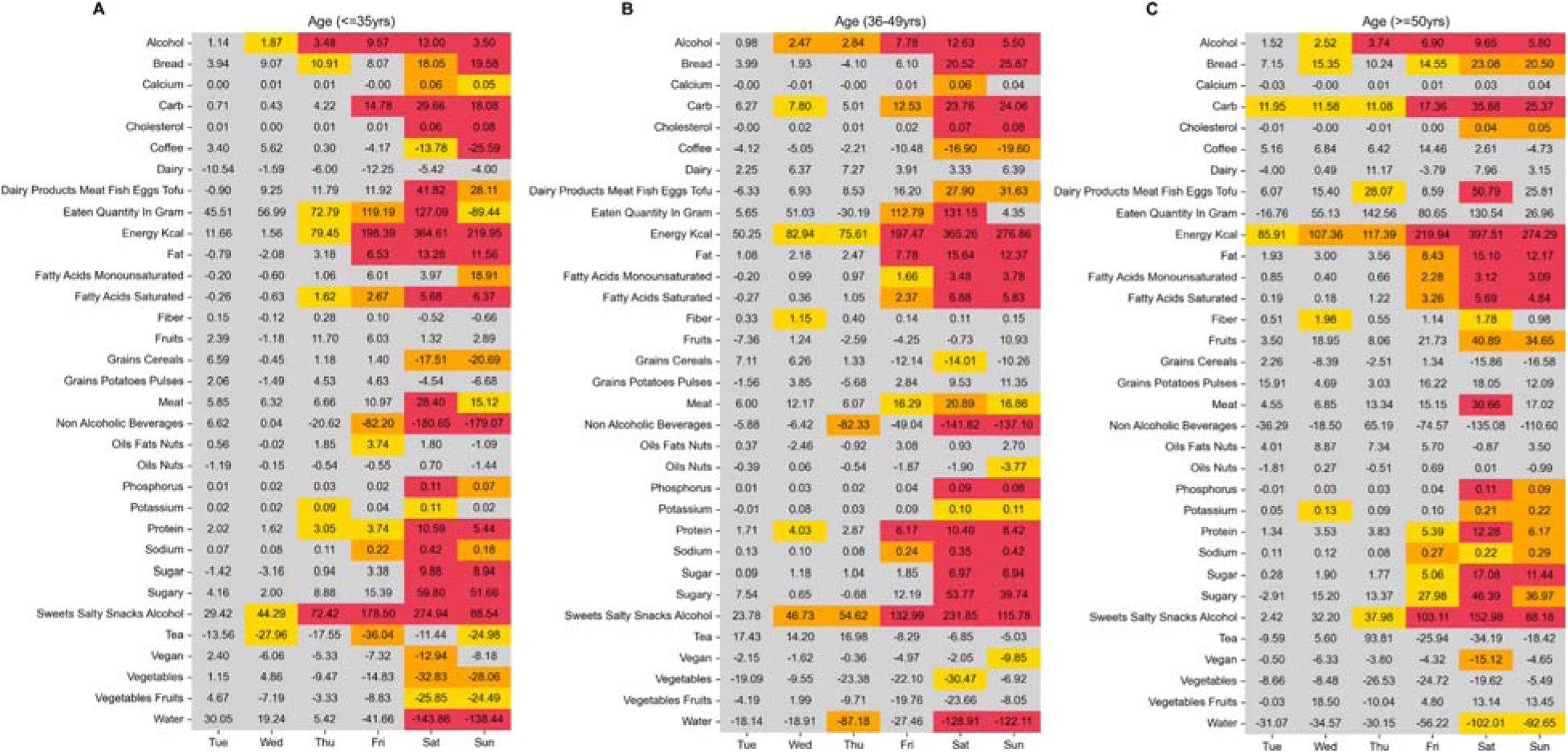
Coefficients and p-values of Linear Mixed Models for various nutrients across different days of the week of the “Food & You” participants, subsetted by age groups. The x-axis represents the days of the week (Tuesday to Sunday), with Monday as the reference day. The cells display the coefficients, indicating the change in nutrient intake compared to Monday. The color of the cells represents the statistical significance of the coefficients: red for p < 0.001, orange for p < 0.01, yellow for p < 0.05, and gray for p ≥ 0.05.

**Supplementary Figure 2:**
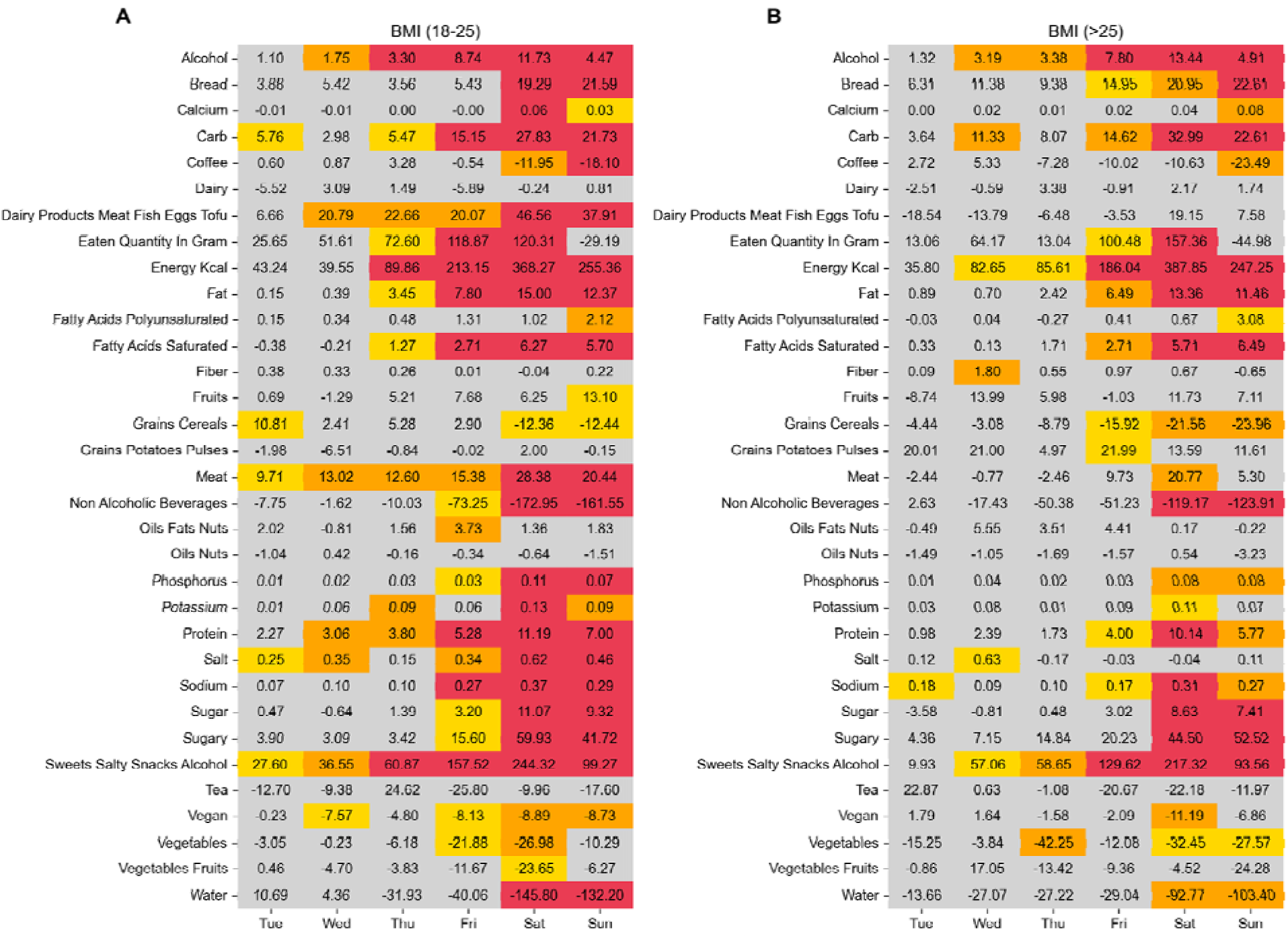
Coefficients and p-values of Linear Mixed Models for various nutrients across different days of the week of the “Food & You” participants, subsetted by BMI. The x-axis represents the days of the week (Tuesday to Sunday), with Monday as the reference day. The cells display the coefficients, indicating the change in nutrient intake compared to Monday. The color of the cells represents the statistical significance of the coefficients: red for p < 0.001, orange for p < 0.01, yellow for p < 0.05, and gray for p ≥ 0.05.

**Supplementary Figure 3:**
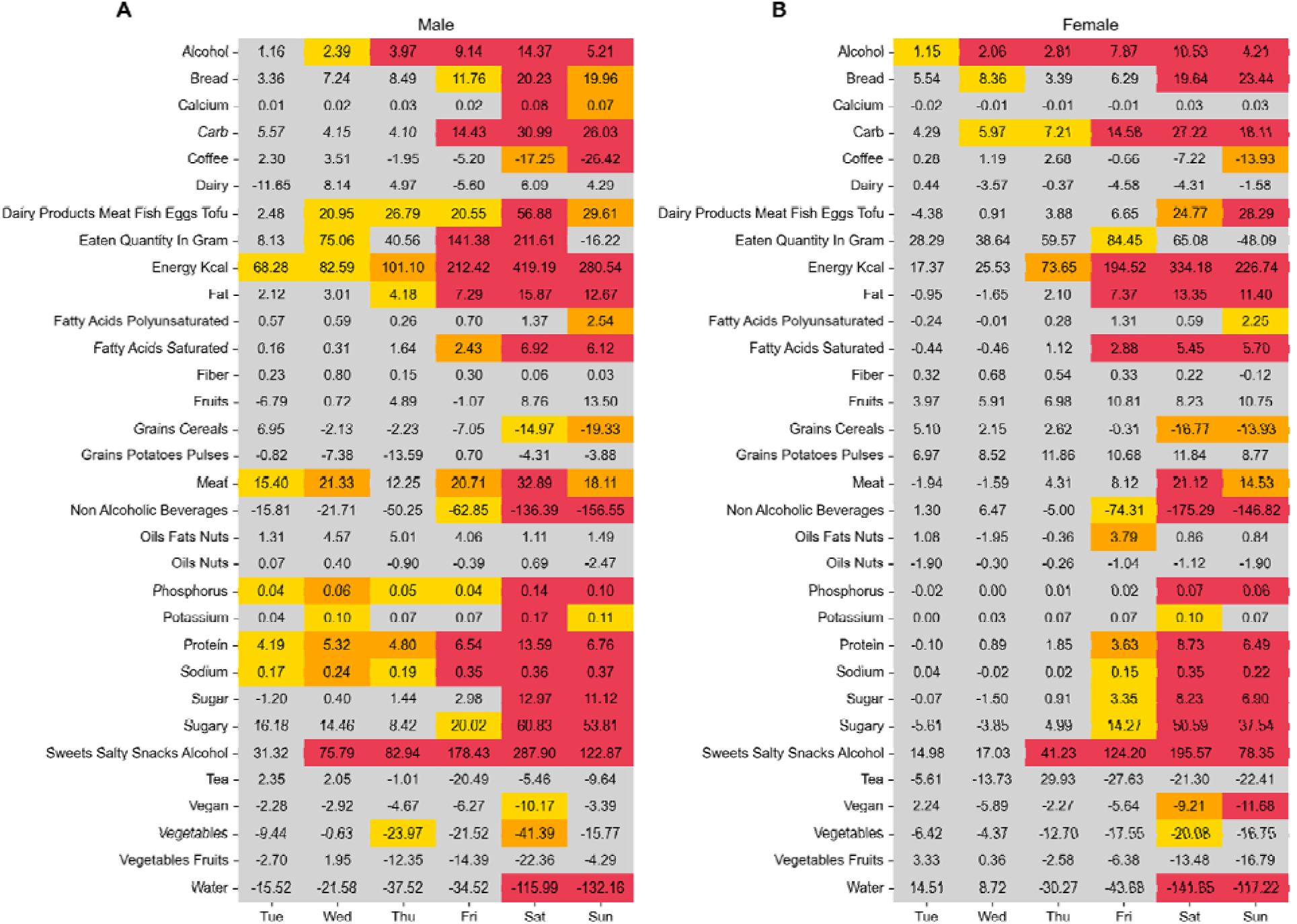
Coefficients and p-values of Linear Mixed Models for various nutrients across different days of the week of the “Food & You” participants, subsetted by gender. The x-axis represents the days of the week (Tuesday to Sunday), with Monday as the reference day. The cells display the coefficients, indicating the change in nutrient intake compared to Monday. The color of the cells represents the statistical significance of the coefficients: red for p < 0.001, orange for p < 0.01, yellow for p < 0.05, and gray for p ≥ 0.05.

**Supplementary Figure 4:**
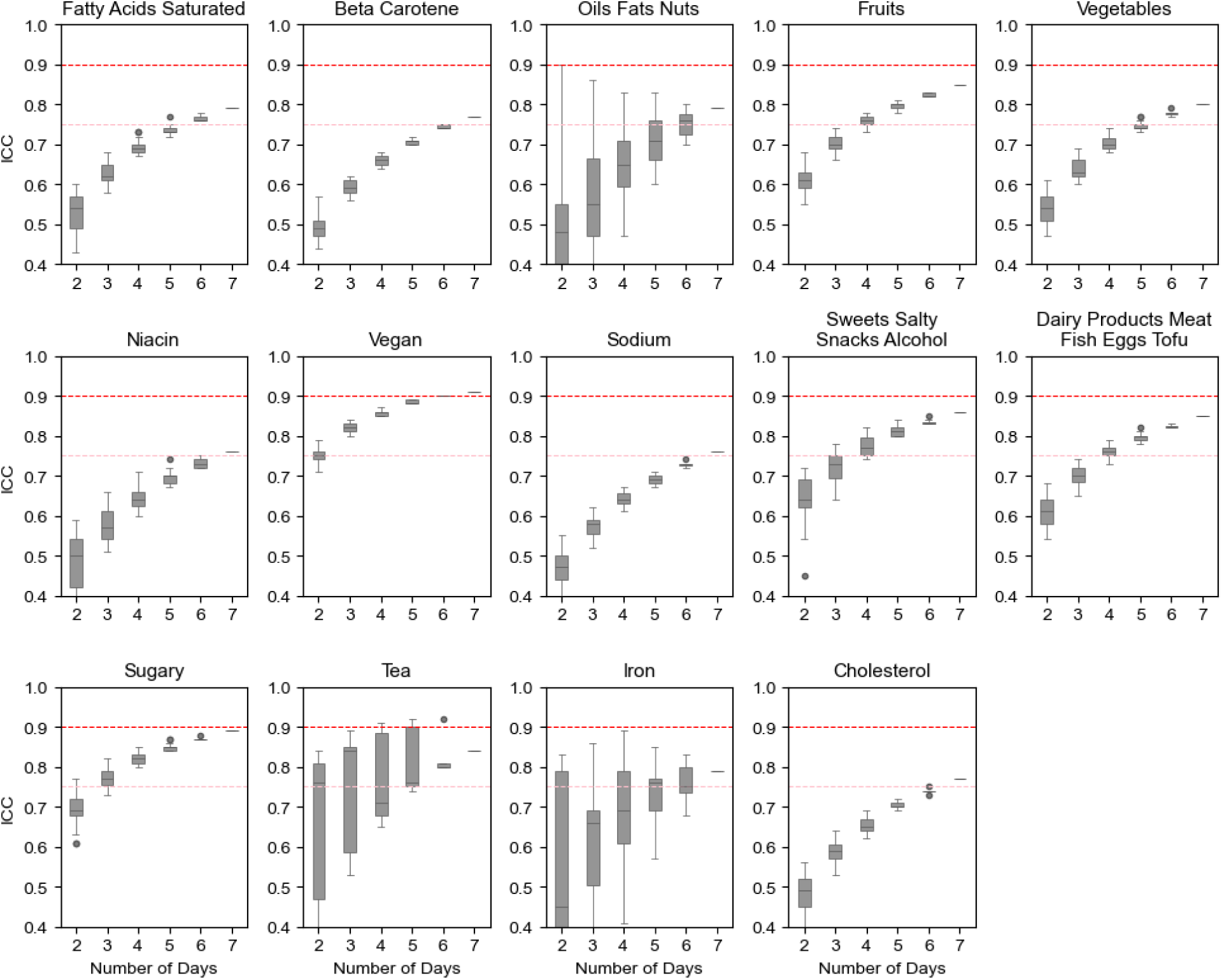
Distribution of Intraclass Correlation Coefficient (ICC) values across different numbers of days for various nutrient intakes for which the ICC value for at least a single combination exceeded 0.75. The x-axis represents the number of days, while the y-axis represents the ICC scores. Each subplot corresponds to a different nutrient, with boxplots indicating the ICC scores for combinations of days for each minimum number of days on the x-axis. The horizontal red lines in each subplot denote the ICC threshold of 0.75 (good reliability) and 0.9 (excellent reliability). Boxplot colors represent different nutrient groups: blue for micronutrients, red for food groups, and green for macronutrients. Note that ICC does not generally reach 1 due to inherent within-subject variability.

**Supplementary Figure 5:**
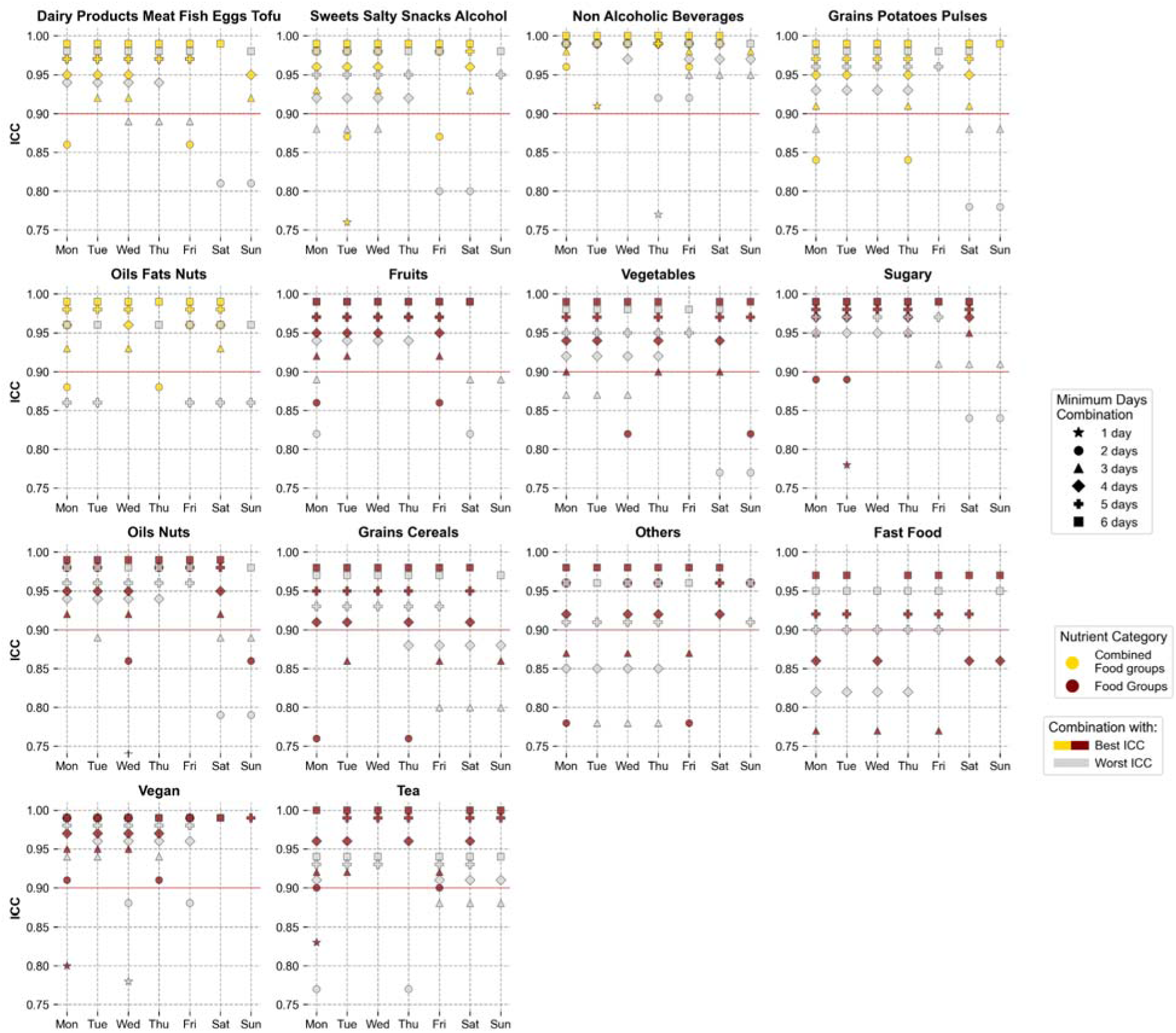
Best and worst day combinations at different minimum days for reliable dietary assessment of different food groups. For each food group and at each number of days, the day combinations which yielded the highest (in color) and lowest ICC scores (in gray) are shown. The plot is ranged between ICC values of 0.75 to 1.0, with the ICC reliability threshold at 0.9 shown as a red line - points lying below this range are not shown.

**Supplementary Figure 6:**
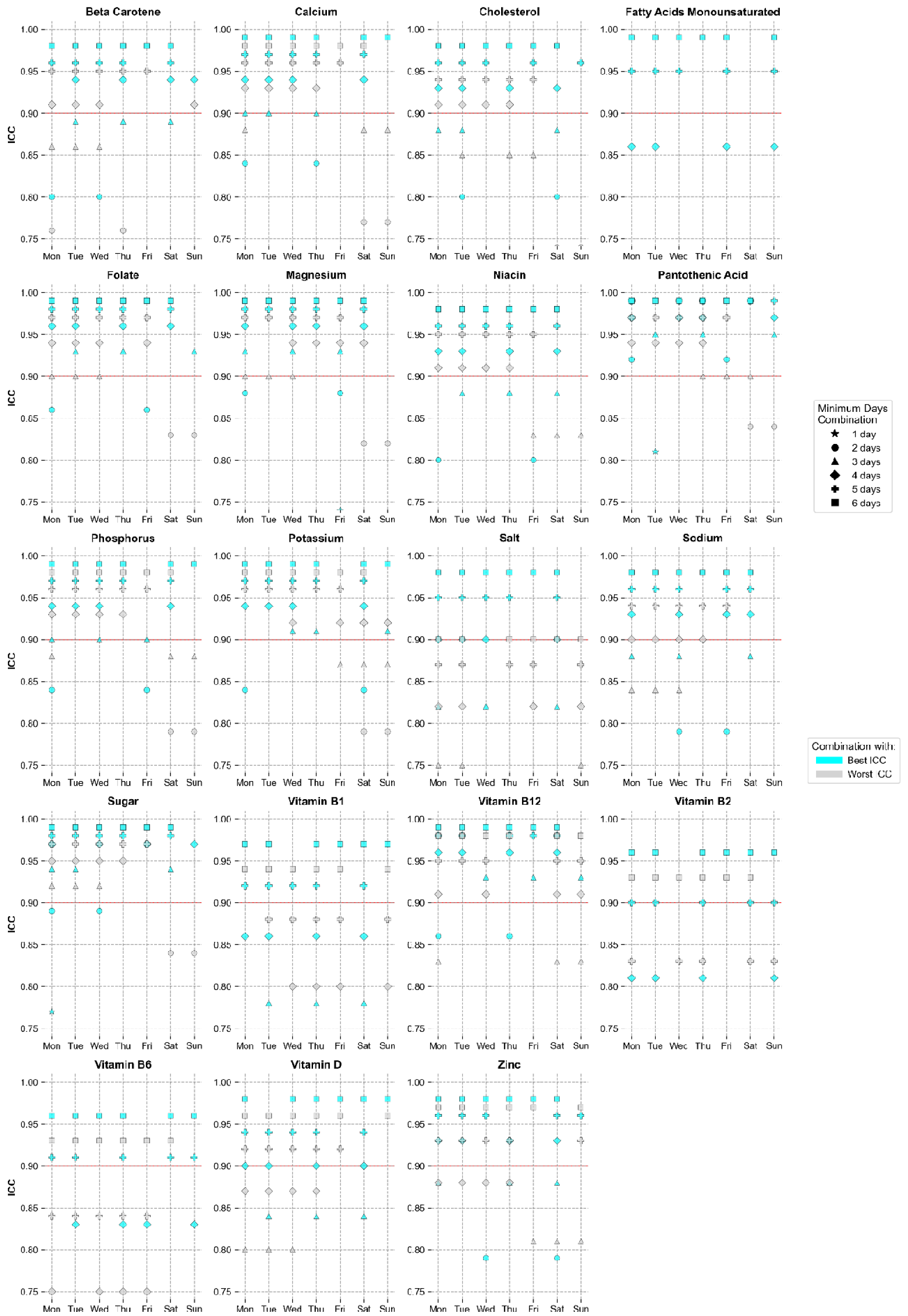
Best and worst day combinations at different minimum days for reliable dietary assessment of different micronutrients. For each nutrient and at each number of days, the day combinations which yielded the highest (in color) and lowest ICC scores (in gray) are shown. The plot is ranged between ICC values of 0.75 to 1.0, with the ICC reliability threshold at 0.9 shown as a red line - points lying below this range are not shown.

